# Forecasting COVID-19 cases: A comparative analysis between Recurrent and Convolutional Neural Networks

**DOI:** 10.1101/2020.11.28.20240259

**Authors:** Khondoker Nazmoon Nabi, Md Toki Tahmid, Abdur Rafi, Muhammad Ehsanul Kader, Md. Asif Haider

## Abstract

When the entire world is waiting restlessly for a safe and effective COVID-19 vaccine that could soon become a reality, numerous countries around the globe are grappling with unprecedented surges of new COVID-19 cases. As the number of new cases is skyrocketing, pandemic fatigue and public apathy towards different intervention strategies are posing new challenges to the government officials to combat the pandemic. Henceforth, it is indispensable for the government officials to understand the future dynamics of COVID-19 flawlessly in order to develop strategic preparedness and resilient response planning. In light of the above circumstances, probable future outbreak scenarios in Brazil, Russia and the United kingdom have been sketched in this study with the help of four deep learning models: long short term memory (LSTM), gated recurrent unit (GRU), convolutional neural network (CNN) and multivariate convolutional neural network (MCNN). In our analysis, CNN algorithm has outperformed other deep learning models in terms of validation accuracy and forecasting consistency. It has been unearthed in our study that CNN can provide robust long term forecasting results in time series analysis due to its capability of essential features learning, distortion invariance and temporal dependence learning. However, the prediction accuracy of LSTM algorithm has been found to be poor as it tries to discover seasonality and periodic intervals from any time series dataset, which were absent in our studied countries. Our study has highlighted the promising validation of using convolutional neural networks instead of recurrent neural networks when it comes to forecasting with very few features and less amount of historical data.

## 1. Introduction

As the northern hemisphere moves into winter, many countries in the world are grappling with sudden surge of COVID-19 cases. As everyone is waiting for a safe, effective and widely available COVID-19 vaccine, new spikes of COVID-19 infections are exacerbating the overall scenario. As the crisis deepens, prudent experts have already warned about the gruesome nature of second wave of COVID-19 as the number of new COVID-19 cases is skyrocketing at unprecedented levels. In the face of second wave of infection, pandemic fatigue and public apathy towards various intervention strategies have prompted people to act impetuously, which are imposing new challenges to the government officials. Different non-pharmaceutical intervention strategies such as wearing of efficacious face coverings, closure of educational institutions, strict travel restrictions and strict containment measures in order to flatten the epidemic curve. In addition, mass level testing and tracing programme are also indispensable for breaking continuous transmission chain. Government officials need to ensure facilitated access to affordable rapid tests. With the help of effective case detection, aggressive contact tracing and local follow-up and support, the probable risks of imminent second wave of COVID-19 in numerous countries can be mitigated.

Different mathematical and statistical paradigms have performed well in predicting the epidemic scenario in different countries since the outbreak of COVID-19. Multiple studies [7, 24, 14, 15, 12, 16] have provided considerable insights to gain a deeper understanding about the transmission dynamics of COVID-19. In the absence of a safe, effective and widely available COVID-19 vaccine, some promising studies [17, 13, 8, 6] have enlightened the importance of different non-pharmaceutical intervention strategies in order to promote effective public health policies in different worst-hit countries when battling against this pandemic. However, it is often really challenging to incorporate all essential real life interactions in a single mathematical model [18, 1] Hence, results are generally based on numerous assumptions and often fail to predict the true outbreak scenario. Nevertheless, a synergy of explicit mathematical models and deep learning networks could be a great way to achieve robust forecasting results in time series analysis [9]. However, this is still an emerging research area which requires more study and experiments.

Deep learning algorithms play a vital role in the analysis and prediction of different epidemic outbreak scenarios. These methods can often provide robust forecasting results which can be used by government officials to deploy appropriate intervention strategies with a view to quelling the spread of any highly transmissible virus. Wang *et al*. used LSTM model in their study [26] to forecast the new COVID-19 cases for 120 days horizon. However, in our study, we have found that LSTM model has failed to capture the right trend with promising accuracy. For instance, it estimated continuous downward trends in Iran, Russia and Peru. However, it is evident that these countries have already encountered with the hit of second wave of COVID-19. In another study [2], Arora *et al*. developed a stacked forecasting model named as Convolutional LSTM” using the concepts of recurrent neural network and convolutional neural network to predict the daily new cases in India. However, that model worked well for a forecasting horizon of only days. This suggests that LSTM architecture does not provide much insights about long term time series forecasting with commendable accuracy. Long term forecasting is undoubtedly a crucial part in terms of policy making and strategic planning. From another point of view, LSTM algorithm works well with large dataset, when it is required to find seasonality in time series analysis. However, with relatively less data points it fails to find any pattern and makes the model too sensitive for forecasting. In such case, a CNN architecture performs well in learning local data patterns and extracting essential features from datasets.

In light of the above concerns, a rigorous comparative study of four deep learning algorithms: long short term memory (LSTM), gated recurrent unit (GRU), convolutional neural network (CNN) and multivariate convolutional neural network (MCNN) has been presented to forecast the number of daily new COVID-19 cases in Brazil, Russia and the UK. Mean absolute percentage error (MAPE) and normalised root mean square error (nRMSE) have been used as performance metrics to evaluate the robustness of model forecasting results. Results based on these metrics suggest that CNN perform better than all other deep learning models. Considering all the scenarios, CNN has turned out to be the best forecasting model due to its power of feature learning. Two deep learning methods: CNN and multivariate CNN have been proposed for forecasting as both methods have shown promising consistency and prediction accuracy in terms of long term forecasting. Importantly, a strong probability of forthcoming second wave of COVID-19 has been unearthed in above-mentioned countries. Robust planning and strategic management can be ensured in these countries based on our forecasting results to avoid disasters in healthcare systems.

The entire chapter is organized as follows. Materials and methods are presented in Section 2. In section 3, forecasting results have been discussed using real-time COVID-19 data of Brazil, Russia and the UK. The paper ends with some insightful experimental results found in this study.

## 2. Materials and methods

In this paper, Deep learning models (CNN, LSTM, GRU, multivariate CNN) are used to forecast new cases using COVID-19 dataset for three countries including Brazil, Russia and the United Kingdom. For Brazil, India, Russia and the United Kingdom data of 291 days are used for training and data of 33 days are used for validation. The forecast is made for 40 days. The performance of the models is evaluated using MSE, nRMSE and MAPE. Models are implemented and trained using Keras module of TensorFlow on Google Collaboratory platform.

### 2.1. Data sources

We have collected the dataset from data up to 18 November, 2020 for Brazil, Russia and the UK from a trusted online repository developed and maintained by Johns Hopkins University [19]. We have extracted the daily new cases and new deaths data for the above-mentioned countries using that repository.

### 2.2. Methods

In this study, specialized four deep learning algorithms: convolutional neural network (CNN), long short term memory (LSTM), gated recurrent unit (GRU) and multivariate convolutional neural network (MCNN) have been applied to understand the future transmission dynamics of COVID-19. Among the mentioned four techniques, LSTM and GRU belong to recurrent neural networks family, whereas CNN and multivariate CNN belong to convolutional neural networks family. As the COVID-19 dataset fluctuates so frequently, moving average method has been used to smoothen our dataset. Dataset for each country was considered as a time series. Our prediction is performed on the moving averaged data. We used rolling mechanism [26] by incorporating windowed data in the time series. By trial-and-error method, we have chosen the best value of window and assigned each country with corresponding best window size. To be precise, window size of days has been used for Russia. Due to high volatility in Brazil’s dataset, window size of 10 days has been used. Moreover, window size of days has been taken for the UK. For COVID-19 time series prediction, recurrent neural networks and their sophisticated variations named LSTM and GRU are widely used. However, the use of convolutional neural network (CNN) in time series analysis is not widespread [10]. In this study, CNN performed very well on both training and validation data as we hypertuned the parameter values. It turns out that CNN worked better than LSTM and GRU on both training and validation data. For forecasting with MCNN, daily confirmed cases and daily deaths have been used to predict the number of newly confirmed cases for the next day. To the best of our knowledge, multivariate convolutional neural network time series analysis is not used before in any study for COVID-19 forecasting. We have listed the parameter values for each model in Tab. 1. We have used root mean squared error (RMSE) as our loss function during the training time. Normalized root mean squared error (nRMSE) and mean average percentage error (MAPE) are used as insightful metrics to evaluate model prediction accuracy and to compare performance between models. We have also incorporated an error boundary for our forecasting zone, where we demonstrated the possible standard deviation from our predicted data.

**Table 1:** Model parameters

| Method | Parameter | Values |
| --- | --- | --- |
| LSTM/GRU | Number of layers | 2 |
|  | Number of units | {50, 20} |
|  | Epochs | 1000 |
|  | Batch size | 300 |
|  | Learning rate | 0.001 |
|  | Optimizer | Adam |
|  | Loss function | MSE |
| CNN/Multivariate CNN | Number of Convolutional layers | 4 |
|  | Number of Units in CNN layers | {32, 64, 128, 256} |
| | Kernel size | $3 \times 3$ |
|  | Number of Dense layers | 2 |
|  | Number of Units in Dense Layers | {10, 1} |
|  | Epochs | 1000 |
|  | Batch size | 300 |
|  | Learning rate | 0.001 |
|  | Optimizer | Adam |
|  | Loss function | MSE |

#### 2.2.1. Moving average

Moving average is one of the most used techniques for understanding trends in time series analysis. It is a way to smooth out fluctuations in time series data and to help distinguish between noise and trends in a dataset. Often in spite of having clear trends and patterns, there might be huge fluctuations found in the dataset, which may cause any forecasting model to perform significantly poor. In that case, the value of any day is replaced by the average of some of its neighboring values. Mathematically, a simple moving average is the mean of the previous n data:

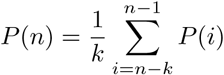

where k is the previous number of days for determining the *i*^*th*^ value. To ensure that the variation in mean corresponds to the variation of data, we have taken the mean of the values of any particular date, the previous two days and the next two days to calculate the moving averaged value of that day Fig. 1. Mathematically,

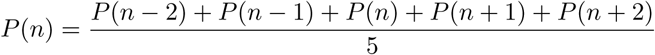

**Figure 1:**
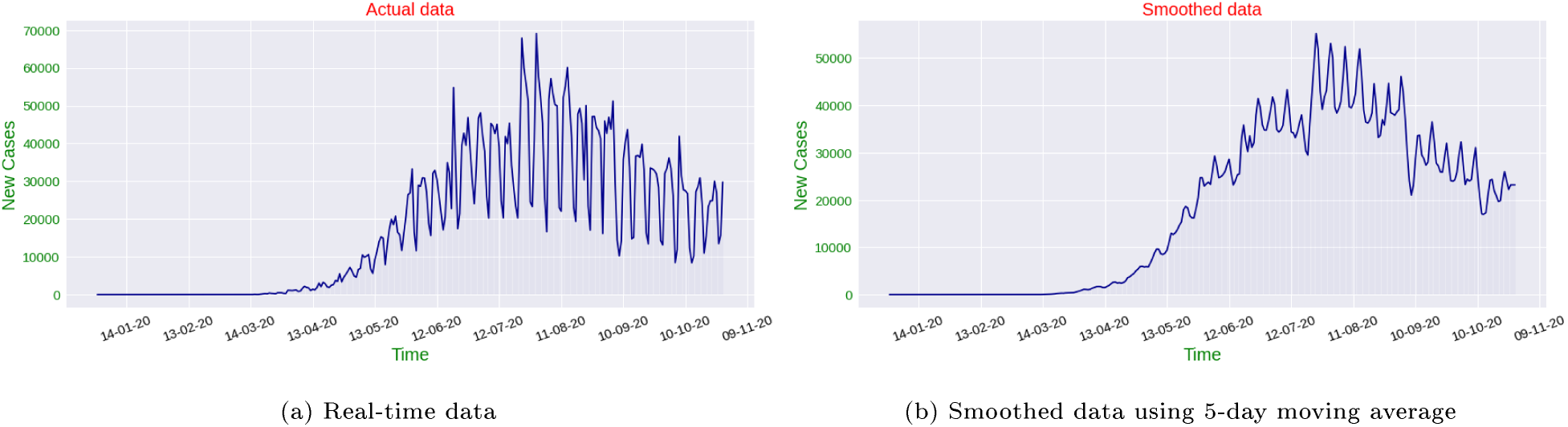
Comparative scenario analysis between real-time data and smoothed real-time data

Here, *P* (*n*) is the value for *n*^*th*^ day.

### 2.3. Deep learning methods

#### 2.3.1. convolutional neural network (CNN)

Convolutional neural network (CNN) is a more generalized version of multilayer perceptron [11]. However, CNN architecture causes over-fitting in the model. Each neuron in the one convolutional layer is connected only to neurons located within a small rectangle in the previous layer. This architecture allows the network to concentrate on low-level features in the first hidden layer Fig.2. then assemble them into higher-level features in the next hidden layer, and so on.

**Figure 2:**
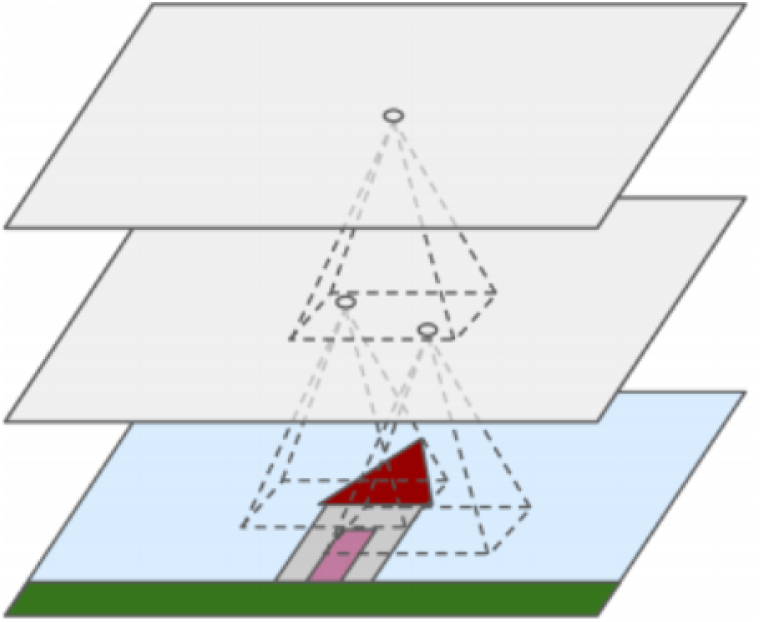
The concept of Convolutional Neural Network: Shrinking down data dimensions to extract most relevant features

To use CNN in time series forecasting Fig.3, four convolutional layers are stacked to extract complex feature and patterns from the time series. Finally, one flatten layer and two dense layers are used to produce the necessary output.

**Figure 3:**
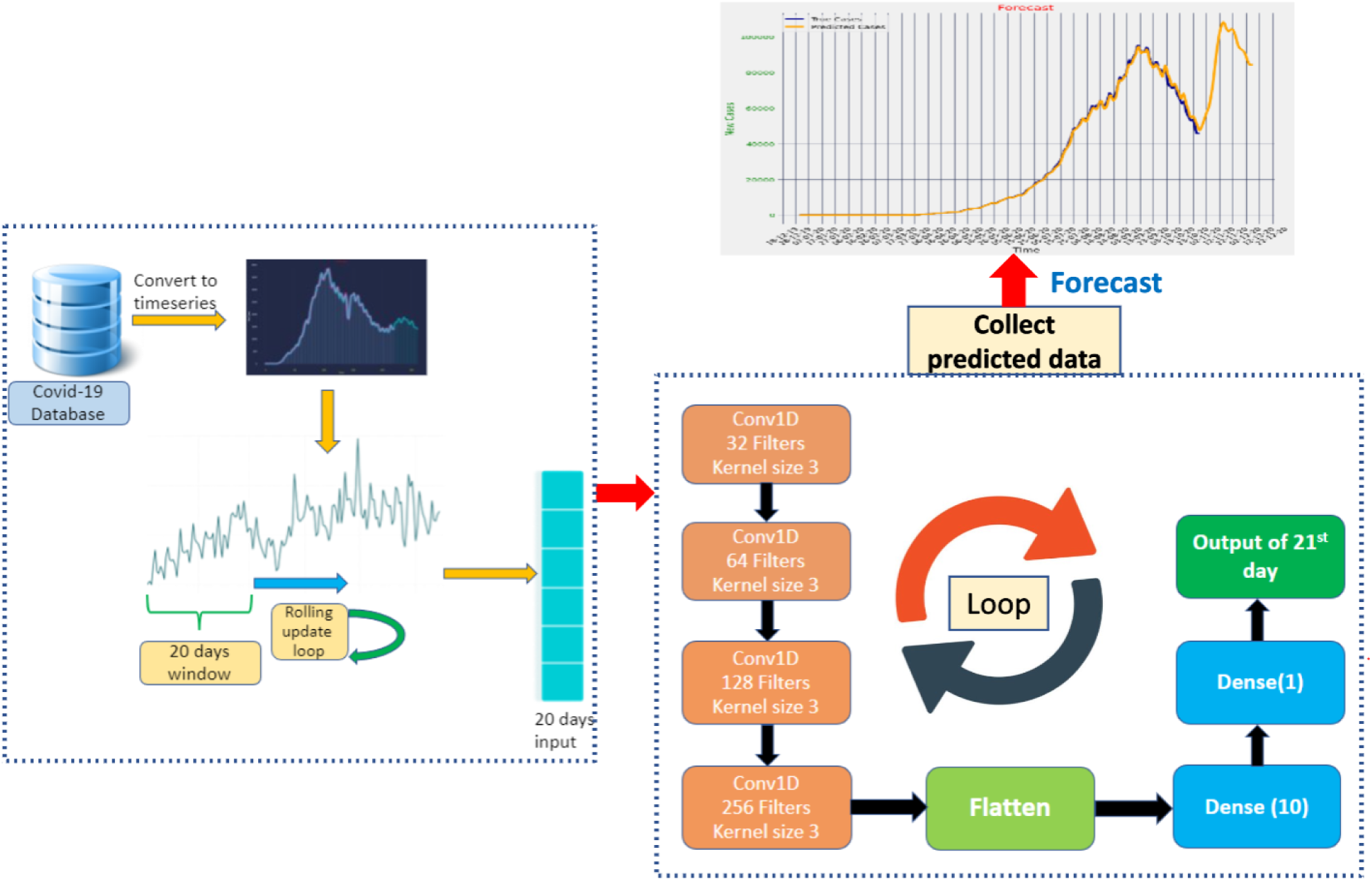
Convolutional Neural Network architecture used in this study

#### 2.3.2. Long short term memory (LSTM)

As traditional Artificial Neural Networks fail to provide desired accuracy while working with sequential data, Recurrent Neural Networks (RNN) or neural networks with loops come into play by maintaining a vector of activation for each time-step and carrying them forward Fig.4. Although RNNs are much better in handling sequential data by connecting previous information to the present inputs, often there might be problems while training deeper models. Due to RNN’s repetitive nature, parameter matrices related to calculating the gradients, tend to shrink down exponentially during backpropagation, thus making the model training process much slower and less effective. This phenomenon is known as Vanishing Gradient problem. As a result, typical RNNs are found to be a poor option while practically dealing with this long-term dependency. The LSTM or Long Short-Term Memory is a well-known variant of traditional RNNs, that addresses this issue by effectively tackling long-term dependencies.

**Figure 4:**
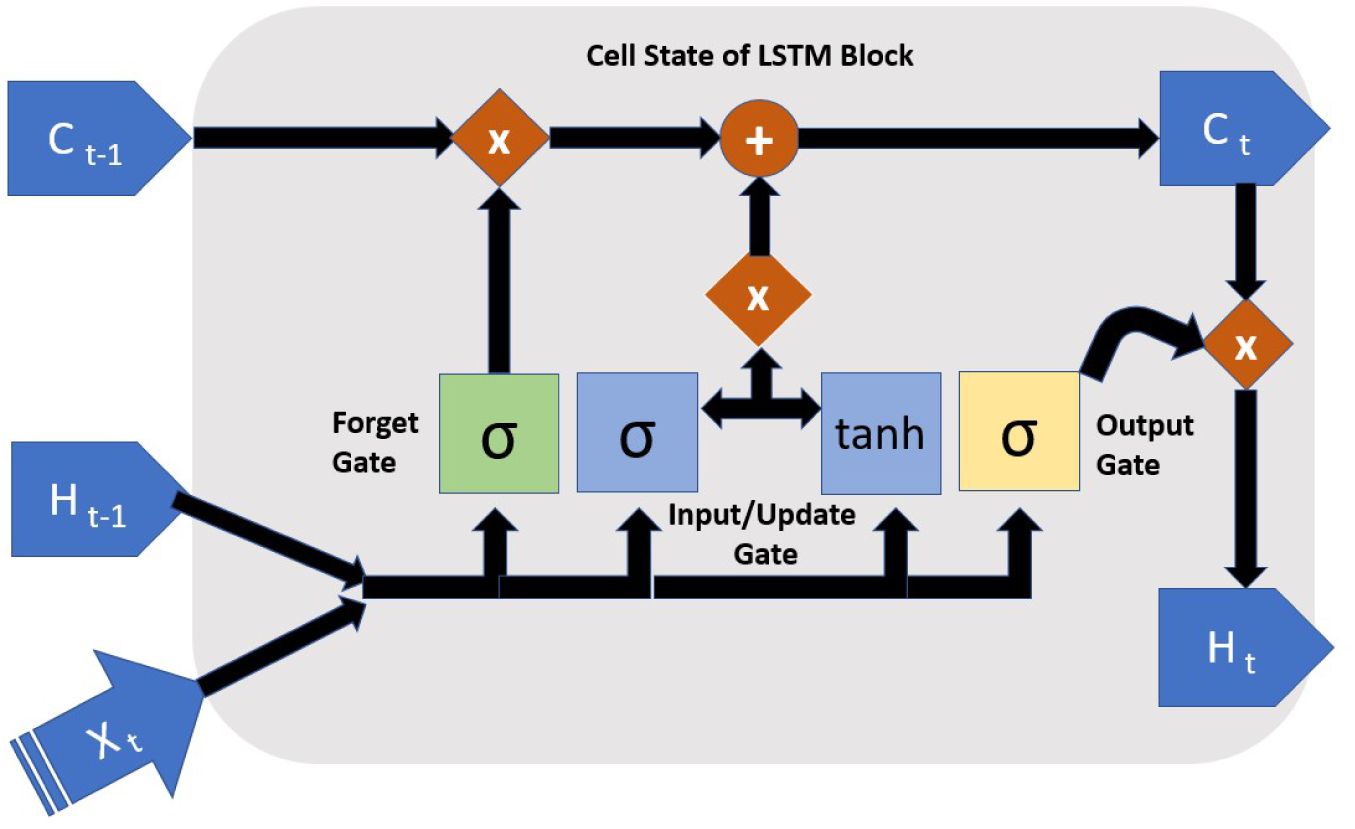
Internal architecture of LSTM network

Structurally, each LSTM block in the network module consists of a set of carefully designed vectorized operations between new inputs and previous outputs, along with the usage of mathematical functions like sigmoid and tanh where necessary, instead of a single layer neural network. In short, LSTM block contains a memory cell and three operational gates with their own weights and bias vectors through which, information pass serially.

First of all, combination of current block input and previous activation values are passed through a nonlinear sigmoid function to filter out which values to store and which values to forget or erase out, as sigmoid function outputs just values in between 0 and 1. This layer is known as the Forget Gate.

Later on, another set of parameters in the Input/Update Gate are used to filter out the same combination of activation from the previous layer and current block input, eventually to be used as the updated input. Then the primary set of input is passed through a tanh (hyperbolic tangent) function to produce new candidate values of the current block. Finally, the linear Vector-Matrix addition of the values from the previous block, scaled by Forget Gate and the current candidate values scaled by the Input/Update Gate is served as the final values of the current block.

Third, the final values of the current block are passed through another tanh activation layer and then scaled by an Output Gate, consisting of another parameterized version of the initial combination of block input and previous activations to produce the newly updated activations of the current block. Thus, using sigmoid activations to decide what to forget and what to store in each block with respect to the previous blocks, LSTM blocks can increase accuracy significantly. Mathematically,

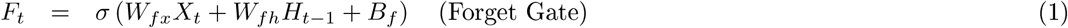

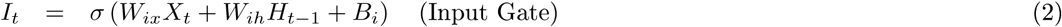

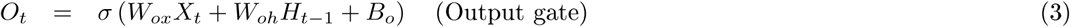

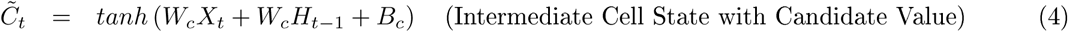

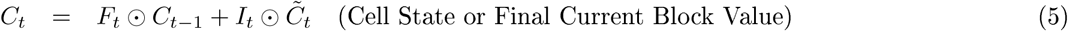

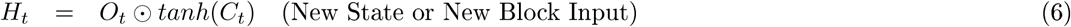

Where, W and B are relevant weights and biases in respective gates associated with each LSTM block, and *X* is the input vector for each block.

#### 2.3.3. Gated Recurrent Unit (GRU)

GRU or Gated Recurrent Unit can be portrayed as a variant of LSTM and it has many similarities with LSTM [5]. Like LSTM, GRU are also primarily used to solve the Vanishing Gradient” problem found in typical RNNs and thus improving the learning of long-term dependencies in the network. GRU blocks also use tanh and sigmoid functions to compute necessary values. But unlike a LSTM block, a GRU block doesn’t have separate memory cells. This type of block also doesn’t have separate Forget gate and the Input/Update gate is responsible for controlling the flow of information. Because of these two structural differences with LSTM, it has less parameters and the design of it is less complex, which eventually makes it more computationally efficient and easier to train. tanh and sigmoid functions are used inside to compute necessary values.

Other than Update gate, GRU blocks have Reset gate. Inside a GRU block, four values are computed, these are - Update gate, Reset gate, candidate activation and output activation Fig. 5. Each gate and candidate activation have their separate weights and biases. Along with their weights and biases, current block input and previous activation values are used as input to compute these three values. In the first step, sigmoid function is used to calculate values of the gates.

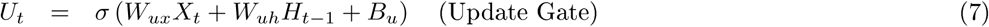

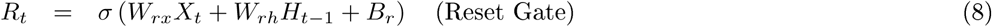

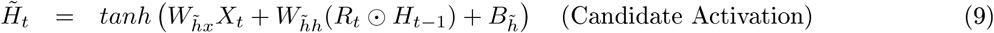

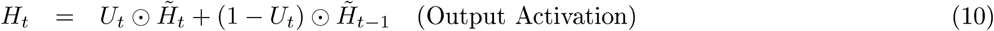

**Figure 5:**
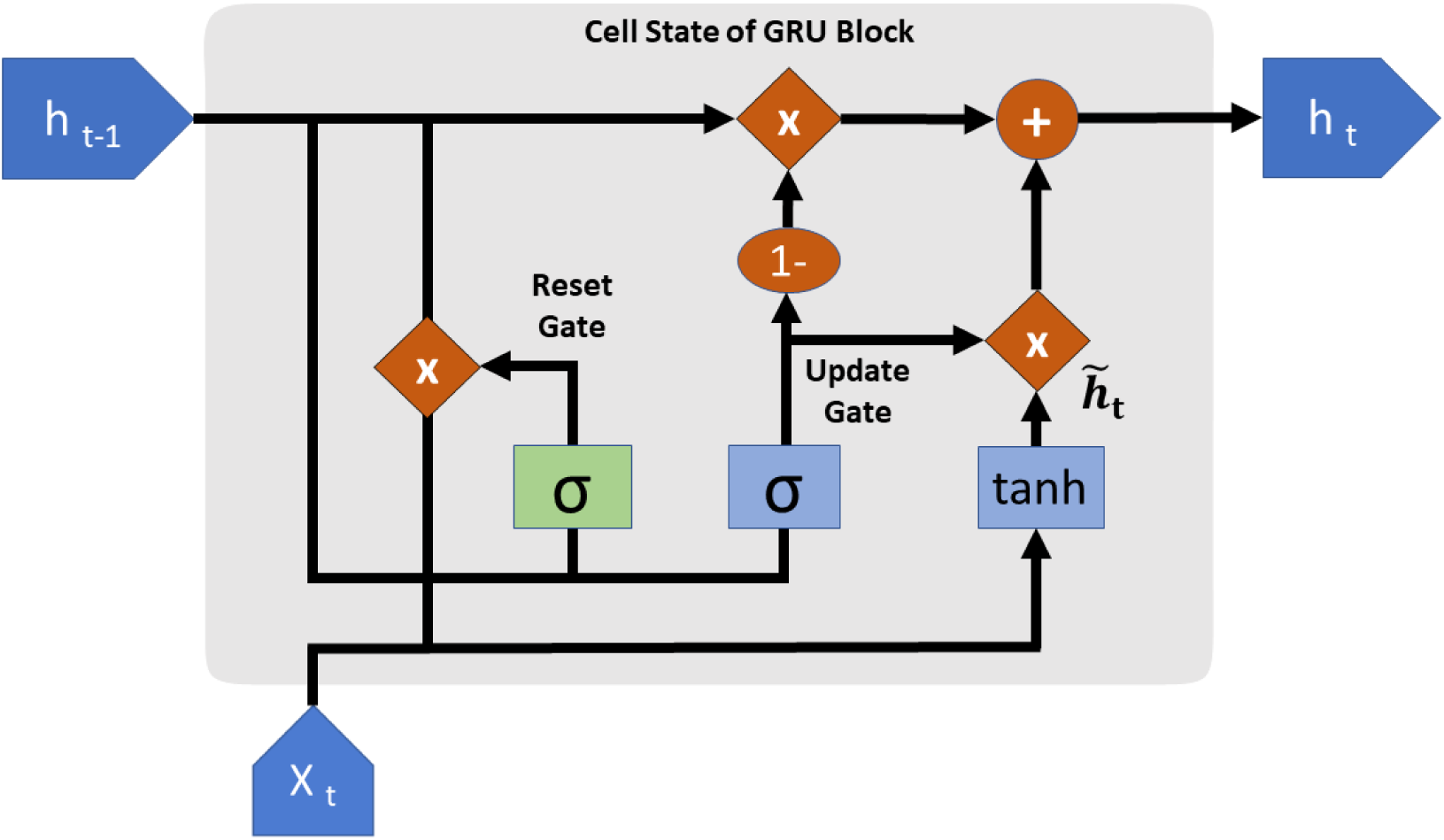
Internal architecture of GRU network

Then, candidate activation is calculated by applying tanh function to a combination of input of Reset gate value and previously mentioned inputs.

Finally, the linear vector addition of the values of candidate activation, scaled by Update gate and previous activation values scaled by the result of subtraction between vector of ones and Update gate produces the values of output activation.

#### 2.3.4. Multivariate convolutional Neural Network

In multivariate time series analysis, multiple features are provided as an input for any particular timestep [3].Along with the targeted feature, this method also predicts the values for other features used as an input. In this study, we have used daily confirmed COVID-19 cases and daily confirmed COVID-19 deaths as our multivariate features as we have observed strong correlation between their trends Fig.6. LSTM networks have previously been used for COVID-19 prediction using multivariate time series analysis [21]. However, we are the first to implement CNN models for multivariate COVID-19 forecasting which provided us with a promising result Tab.2.

**Table 2:** Values of evaluation metric from various deep learning models on different countries

| Country | Deep learning model | Evaluation metric values |  |  |  |
| --- | --- | --- | --- | --- | --- |
| | | $nRMSE_{train}$ | $nRMSE_{val}$ | $MAPE_{train}$ | $MAPE_{val}$ |
| Brazil | LSTM | 0.16 | 0.197 | 14.92% | 13.33% |
|  | GRU | 0.103 | 0.124 | 8.39% | 10.267% |
|  | CNN | 0.054 | 0.086 | 8.20% | 6.94% |
|  | Multivariate CNN | 0.068 | 0.109 | 16.73% | 8.21% |
| Russia | LSTM | 0.203 | 0.05 | 14.03% | 4.64% |
|  | GRU | 0.05 | 0.01 | 5.76% | 1.199% |
|  | CNN | 0.06 | 0.014 | 6.61% | 0.85% |
|  | Multivariate CNN | 0.05 | 0.01 | 4.79% | 0.86% |
| United Kingdom | LSTM | 0.26 | 0.065 | 16.36% | 5.25% |
|  | GRU | 0.11 | 0.036 | 7.0% | 2.997% |
|  | CNN | 0.12 | 0.048 | 12.0% | 3.75% |
|  | Multivariate CNN | 0.15 | 0.094 | 14.48% | 7.797% |

**Figure 6:**
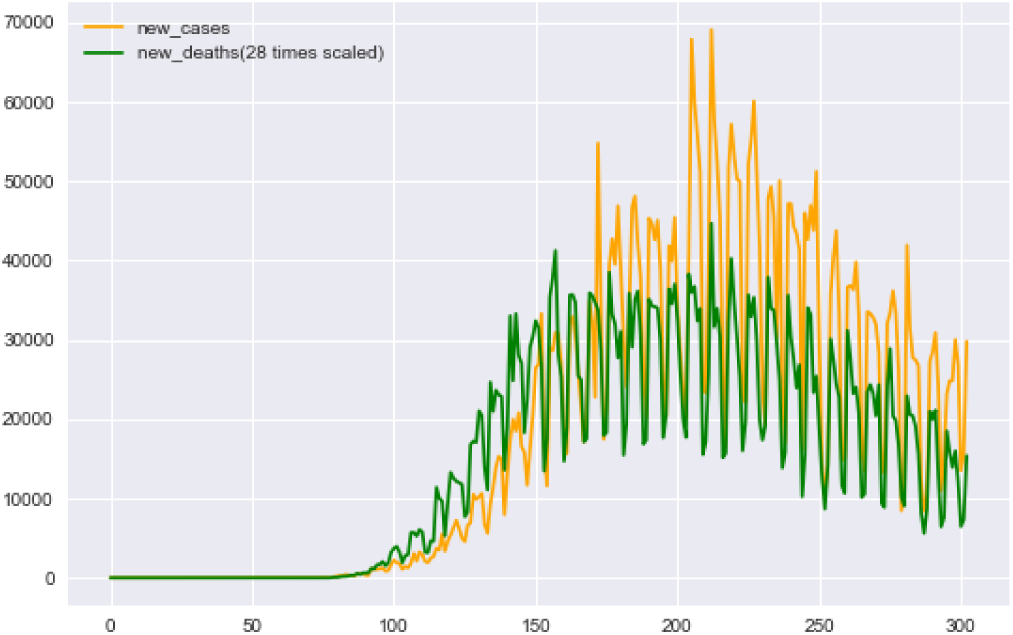
Correlation between new cases and new deaths. Here the ‘new_deaths’ data has been scaled 28 times to visualize the trend more clearly.

Normalized root mean squared error (nRMSE) and mean absolute percentage error (MAPE) have been implemented to understand relative performance of different deep learning models. Following are the formulas to determine the values of these metrics:

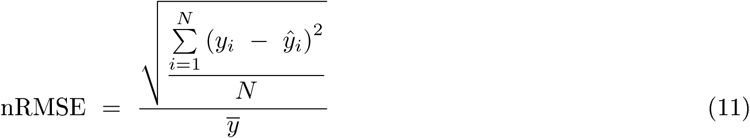

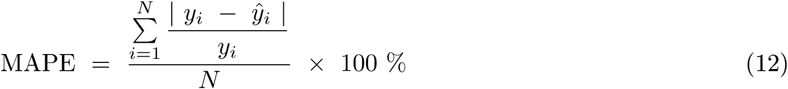

Here, *y*_*i*_ are the actual values, *ŷ*_*i*_ are the values predicted by the model and N is the number of observations. In the nRMSE formula, 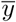 is the mean of all the predicted values. As the difference between actual values and predicted values are squared in nRMSE and absolute values are taken in MAPE, the problem of positive and negative errors canceling each other out is avoided.

## 3. Results

In this analysis, we have used four deep learning architectures to forecast the new cases of Brazil, Russia and the United Kingdom for next 40 days. We have summarized the values of normalized root mean squared error train (*nRMSE*_*train*_), normalized root mean squared error validation (*nRMSE*_*val*_), mean average percentage error train (*MAPE*_*train*_), mean average percentage error validation (*MAPE*_*val*_) in Tab.2. Along with new COVID-19 cases, we have also implemented our best validated CNN model to forecast new death cases for above countries with a forecasting horizon of 40 days.

In our study we have found that CNN performed the best in terms of consistency among all four deep learning models. In this section, study findings are summarized below.

Due to the highly inconsistent daily test numbers, it has been found that Brazil’s daily data is highly volatile and noisy. To address this issue, moving average technique has worked quite well to capture the real trends Fig. 1. Although Brazil had already passed its peak of the first wave with approximately 69, 000 daily cases, our analysis is showing that it has already headed towards another wave. It is evident from Fig. 8.that our CNN model has the lowest metrics value with *nRMSE*_*val*_ = 0.086 and *MAPE*_*val*_ = 6.94%. In addition, the model projected that the country has witnessed another wave by December with a tentative peak of 80, 000 new cases on daily basis illustrated in Fig. 9. Multivariate CNN model was the second best performer in validation data with *nRMSE*_*val*_ = 0.109 and *MAPE*_*val*_ = 8.2% and it also suggests that another wave is approaching in December. It also predicts that the number of cases might decrease soon after reaching the peak of second wave. Two other models: LSTM (*nRMSE*_*val*_ = 0.2 and *MAPE*_*val*_ = 13.33%) and GRU (*nRMSE*_*val*_ = 0.12 and *MAPE*_*val*_ = 10.267%) have also predicted that the number of new cases might increase without showing lucid indication of impending second wave. Moreover, Fig. 7 shows that the tally of daily deaths in Brazil is going to increase gradually until early December although the intensity might not be as high as the first wave with a peak of around 800 new deaths per day.

**Figure 7:**
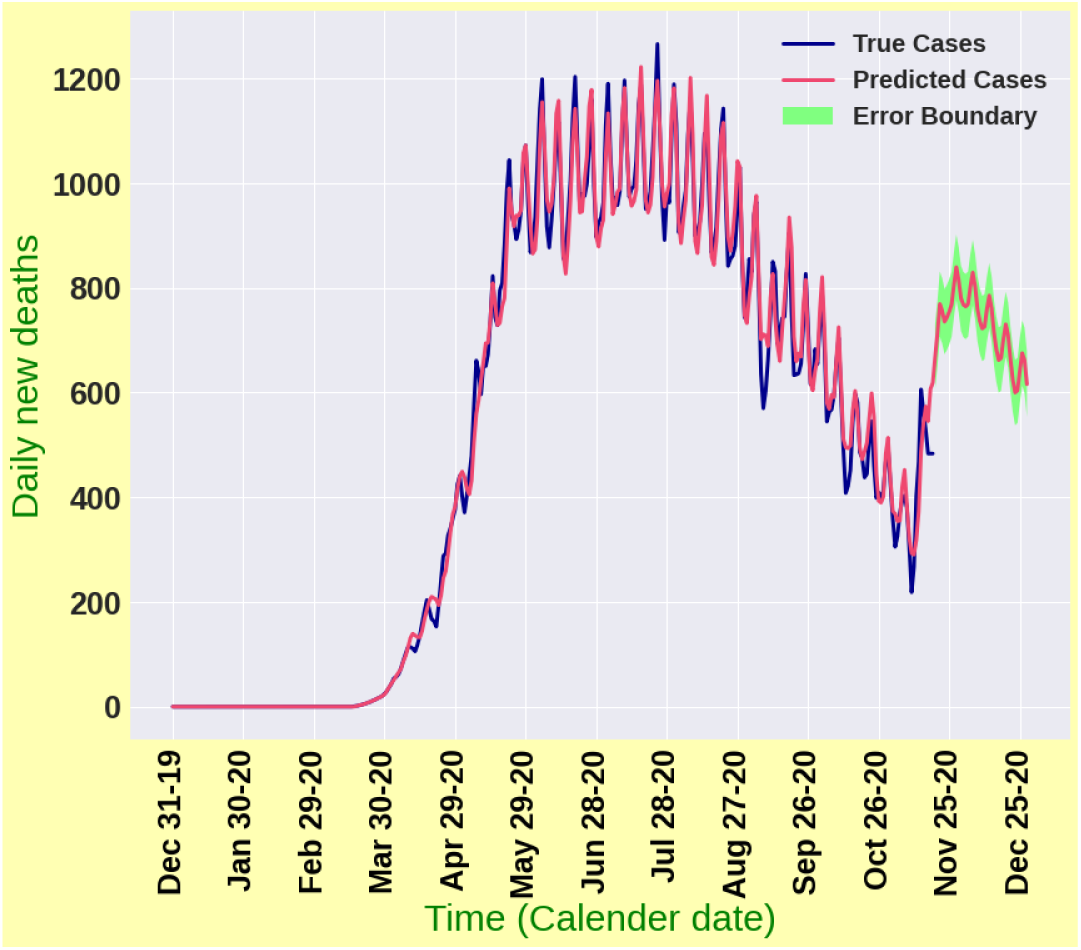
Forecasting results for daily death cases in Brazil

**Figure 8:**
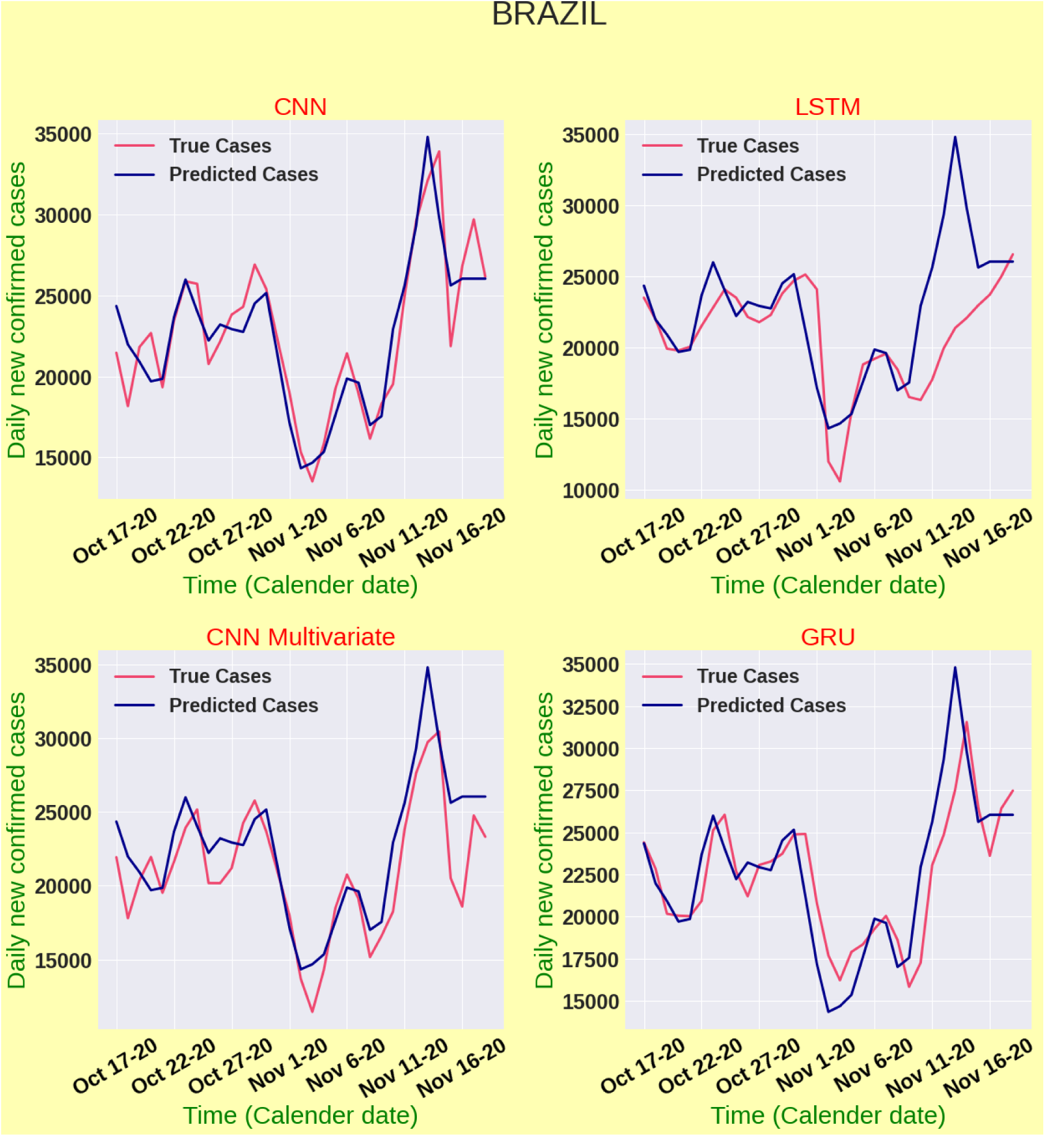
Model validation for Brazil

**Figure 9:**
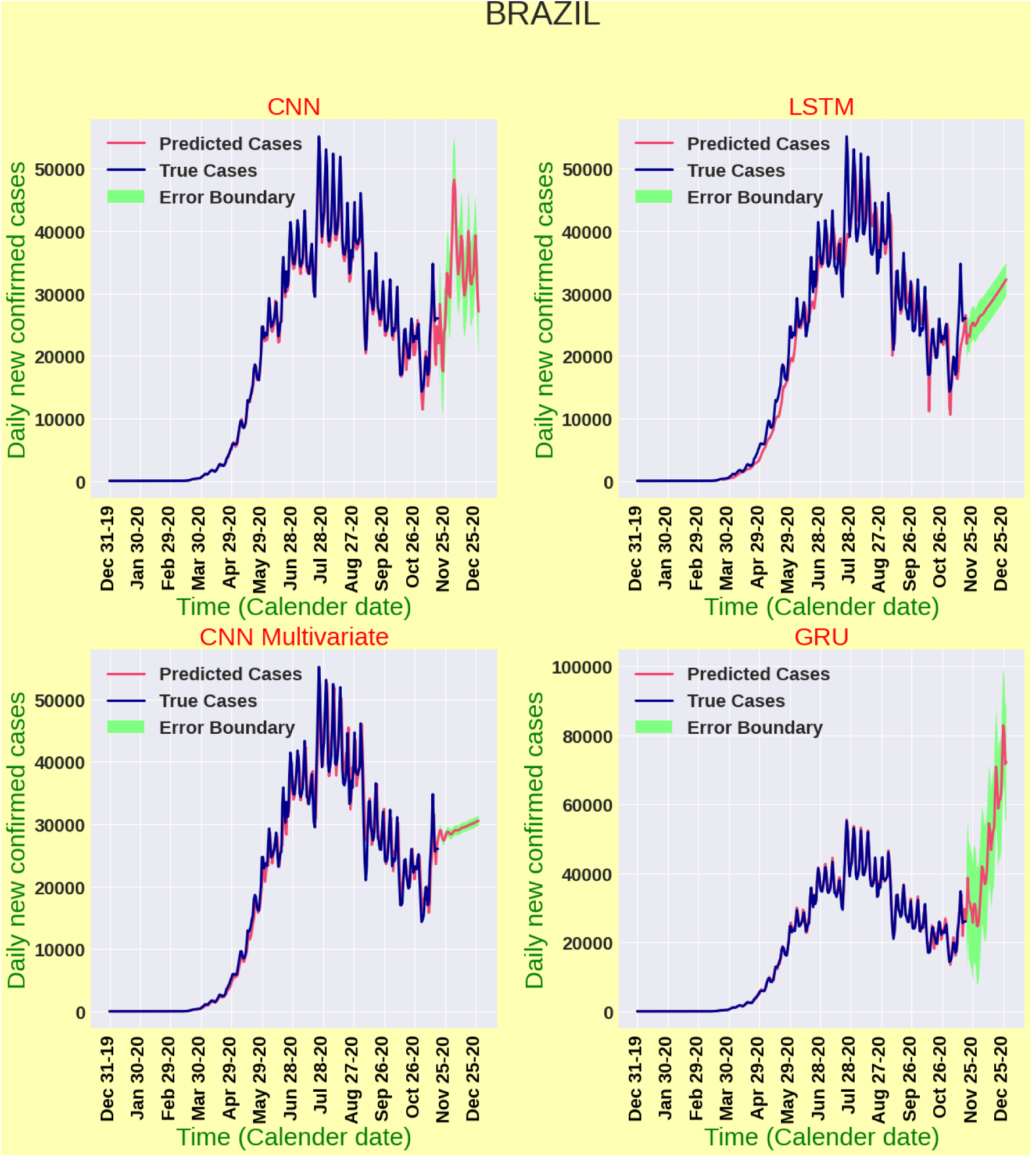
Forecasting results for daily new cases in Brazil

Russia has already gone through their first wave and in midway of second wave. Our model CNN (*nRMSE*_*val*_ = 0.01, *MAPE*_*val*_ = 0.85%) performs the best in terms of validation data for Russia depicted in Fig. 10. It predicts that the number of daily cases might continue to increase and reach nearly 30, 000 by late December. Multivariate CNN (*nRMSE*_*val*_ = 0.01, *MAPE*_*val*_ = 0.86%) shows that the cases might mount to 25, 000 and then might follow a downward trend. In contrast, LSTM (*nRMSE*_*val*_ = 0.05 and *MAPE*_*val*_ = 4.64%) and GRU (*nRMSE*_*val*_ = 0.014 and *MAPE*_*val*_ = 1.2%) indicates a downtrend of cases. Fig. 12 enlightens that the number of daily deaths could skyrocket alarmingly and reach 1500 on daily basis by late December if this trend continues. It is often really challenging to capture the outbreak scenario as it is highly dynamic and depends on deploying various intervention strategies. However, as cases are projected to increase by two of our highly validated models, more precise strategic planning is required in combating against the forthcoming threatening pandemic scenario.

**Figure 10:**
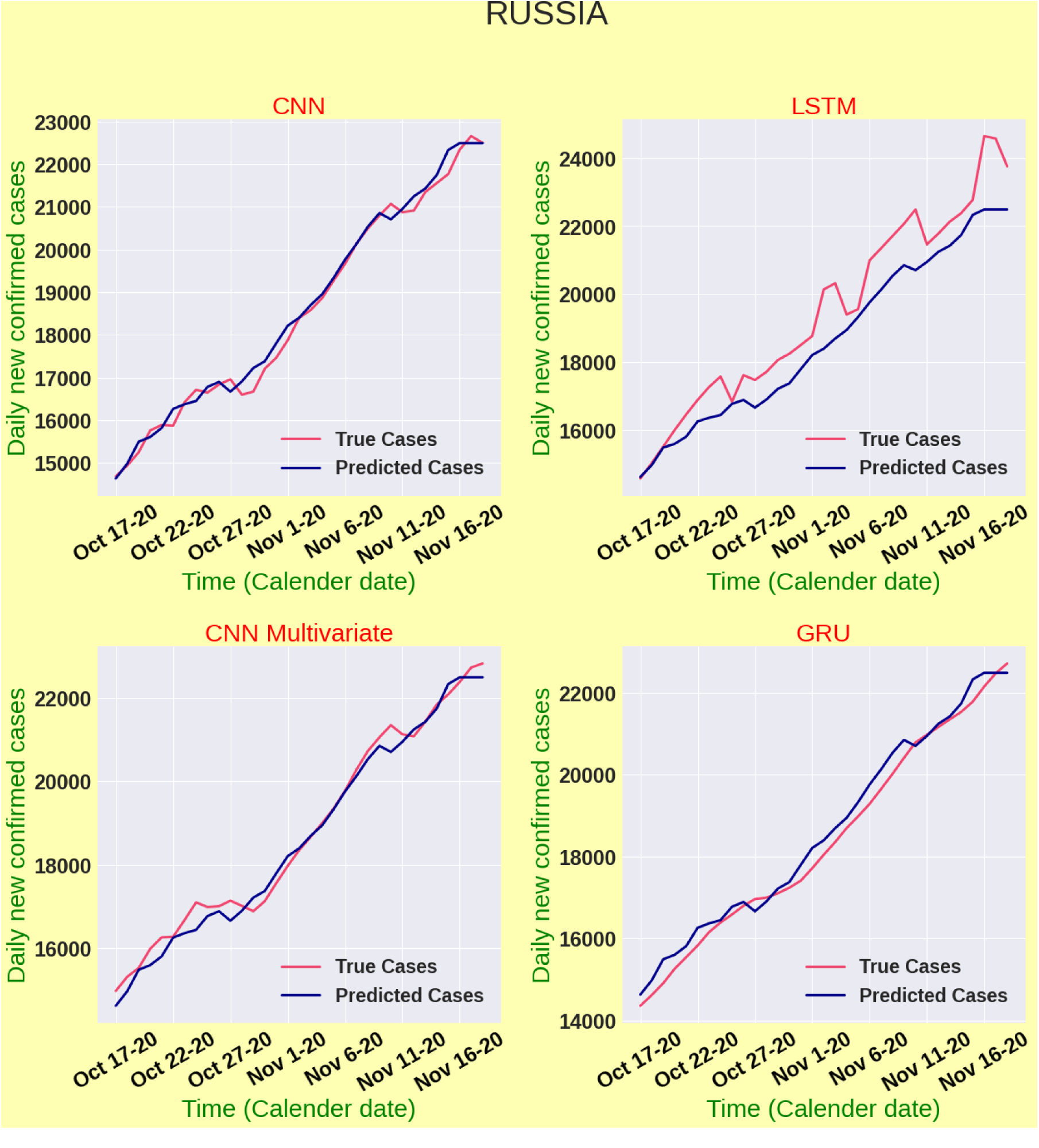
Model validation for Russia

**Figure 11:**
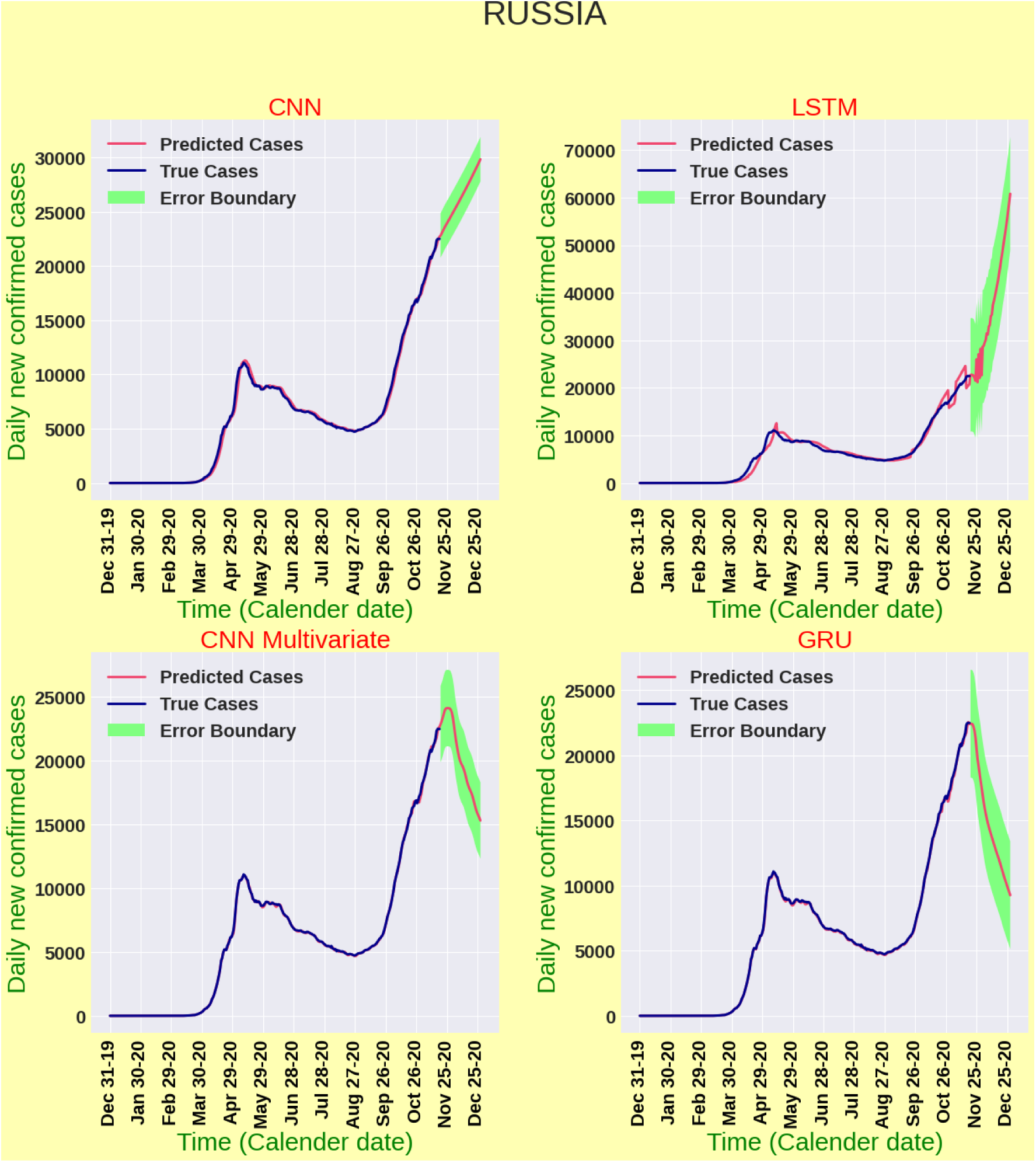
Forecasting results for daily new cases in Russia

**Figure 12:**
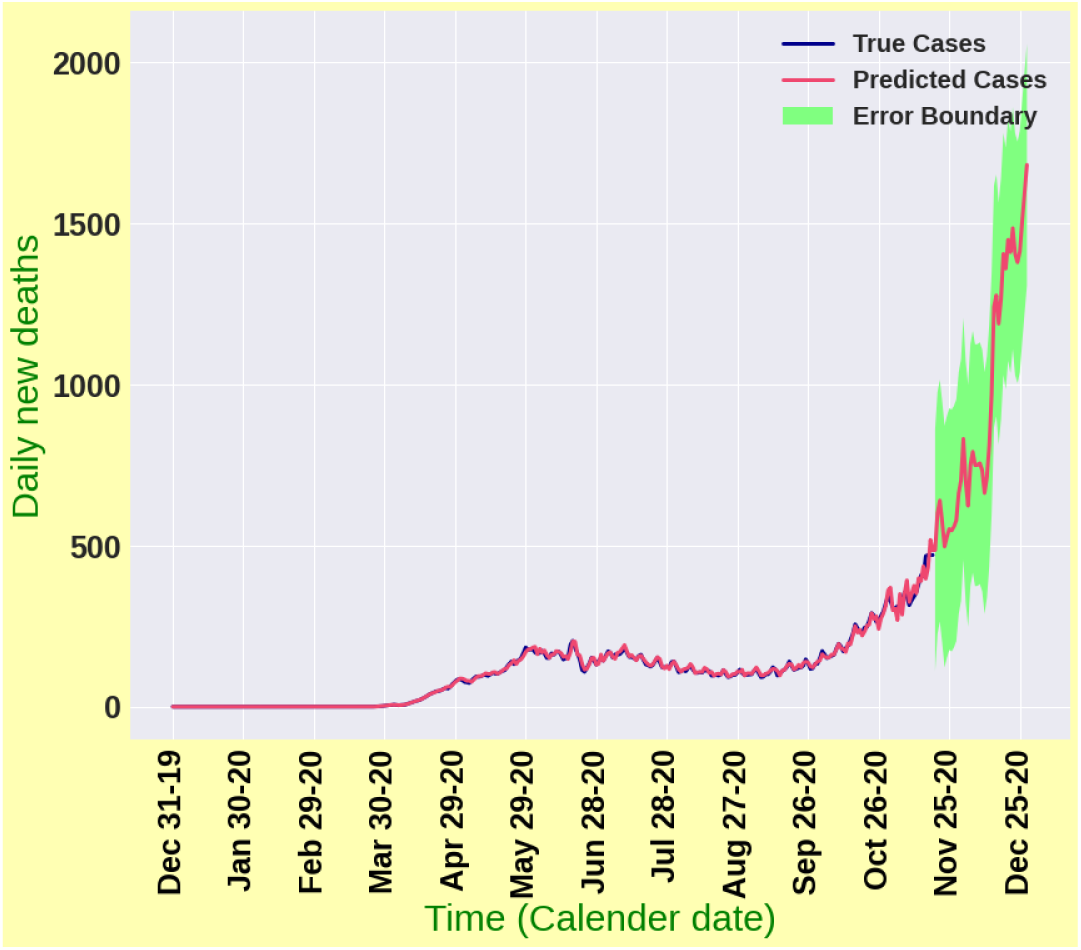
Forecasting results for daily death cases in Russia

The United Kingdom (UK) has already witnessed two waves of COVID-19 illustrated in Fig. 15. In the first wave, the transmission chain was quite well controlled with well implemented and maintained lockdown policies. Moreover, the situation was comparatively better than most of the European countries until September. However, due to restriction fatigue and public apathy towards lockdown, the country is grappling with new surges of cases after September and already witnessed a recent peak of about 33, 000 daily cases. It has been found in our analysis that the number of daily cases may hold the upward trend in upcoming days. Fig. 14 enlightens the fact that, our CNN model achieved an *nRMSE*_*val*_ score of 0.04809 and *MAPE*_*val*_ score of 3.75%, while the Multivariate CNN model shows *nRMSE*_*val*_= 0.09 and *MAPE*_*val*_= 7.8%. On the other hand, *nRMSE*_*val*_ = 0.06481 and *MAPE*_*val*_ = 5.25% have been found for LSTM model. Finally, the GRU model hits *nRMSE*_*val*_ = 0.04 and *MAPE*_*val*_ = 2.997% metric. Comparing with other four models, it has been found that the GRU model best validates the data with the lowest percentage error. The CNN model also performs quite well in this case. However, the CNN and Multivariate CNN models have also projected that the probability of case decline soon after the peak is reached. According to our model projection illustrated in Fig. 13, the number of daily deaths is going to escalate by overshooting the intensity of prior waves in near future and could reach 1000, which could be a great concern for the country’s strategy builders.

**Figure 13:**
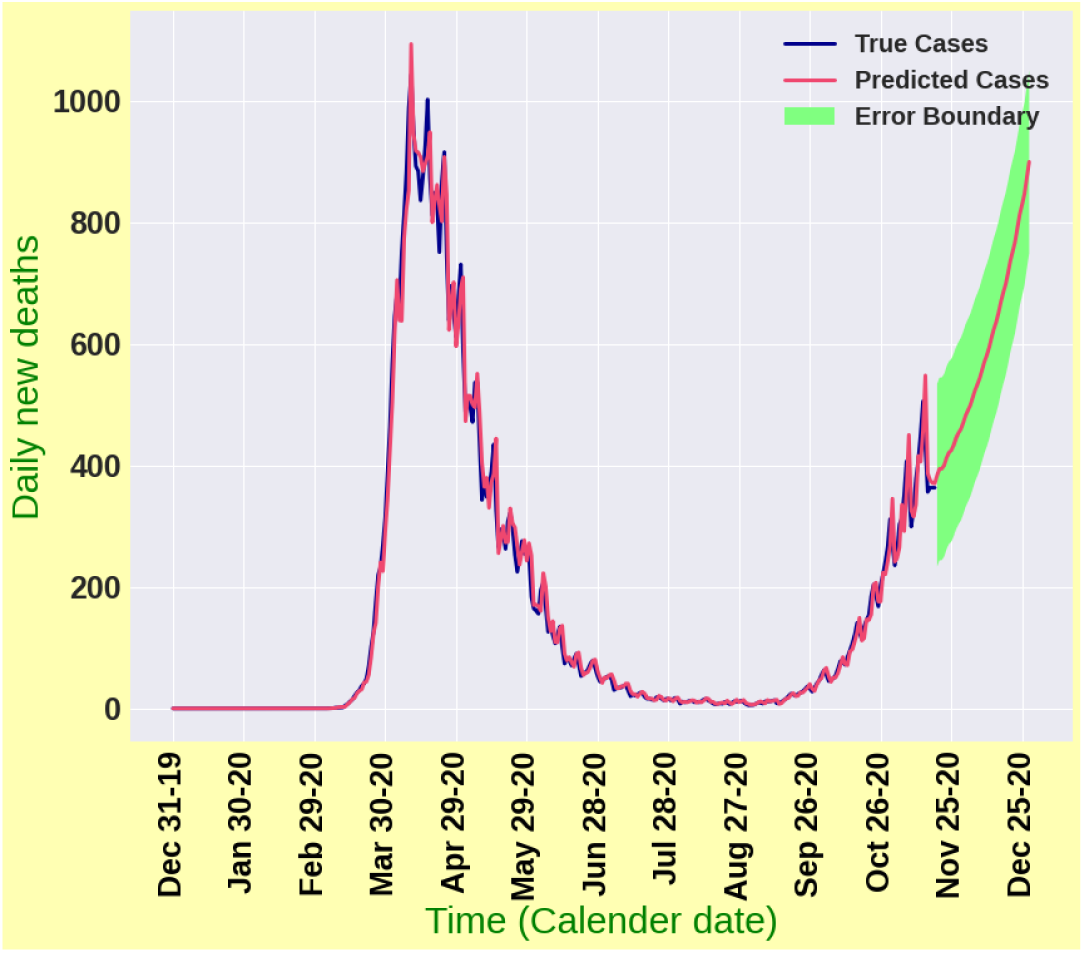
Forecasting results for daily death cases in the United Kingdom

**Figure 14:**
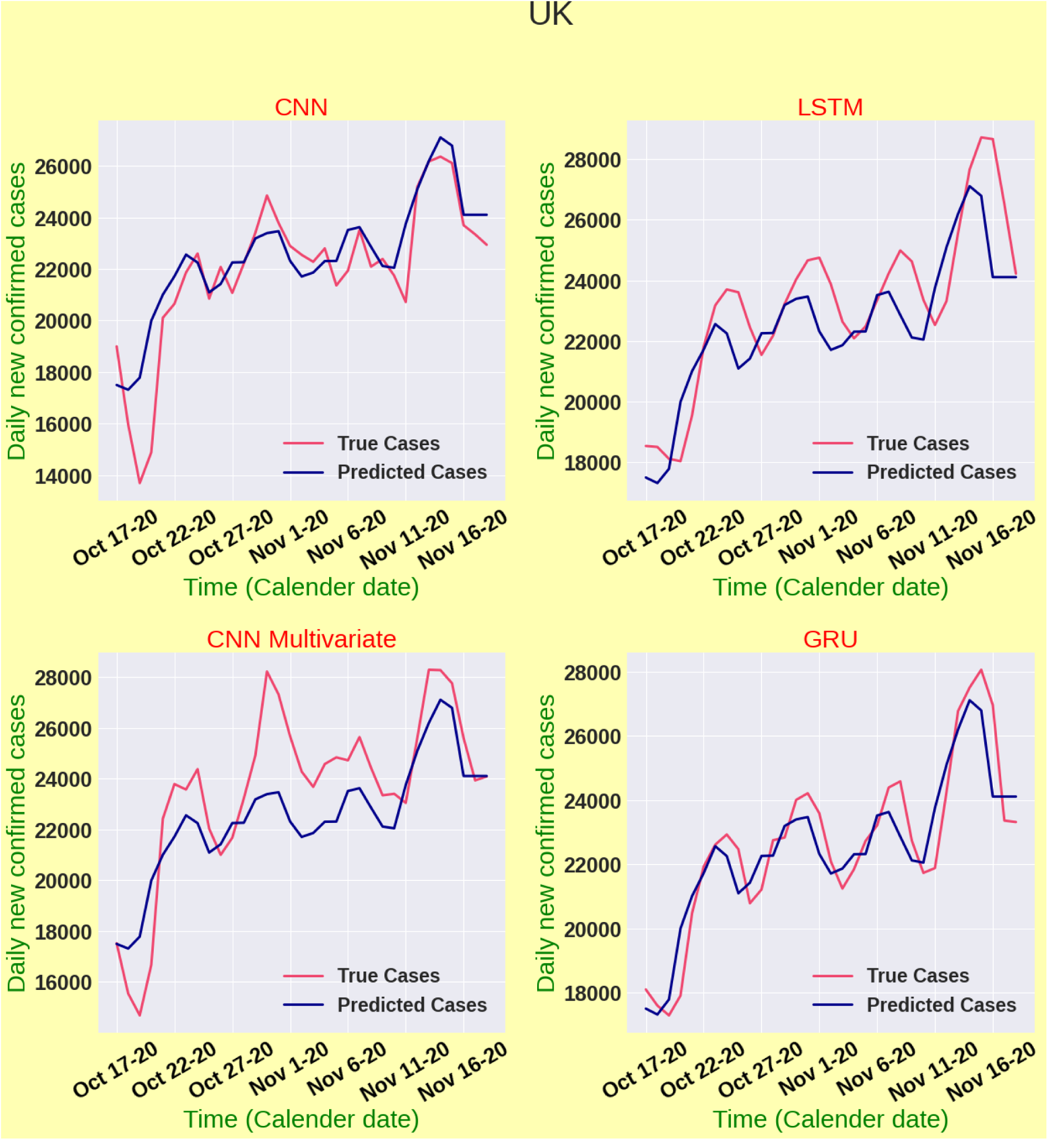
Model validation for the United Kingdom

**Figure 15:**
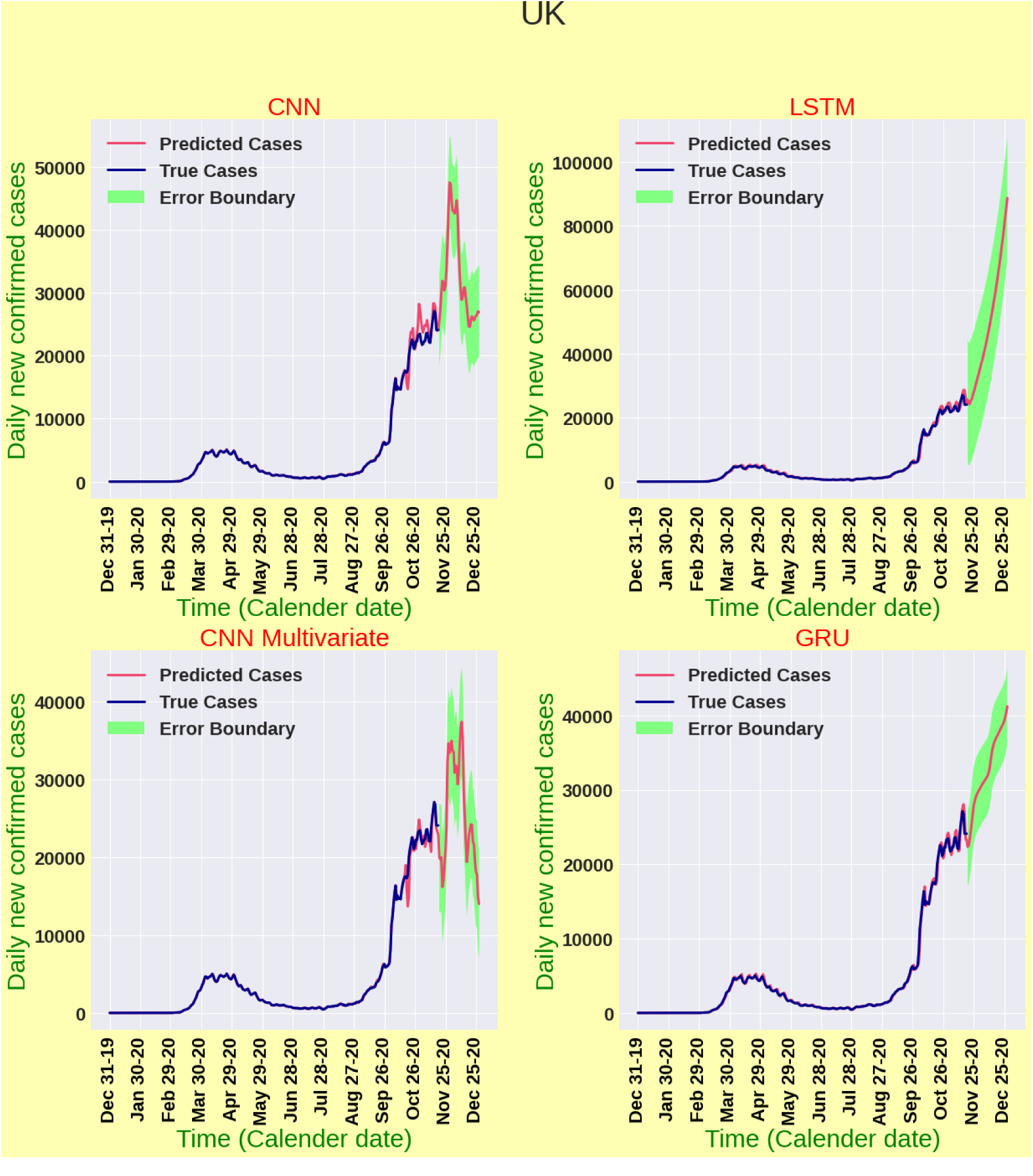
Forecasting results for daily new cases in the United Kingdom

It is worth mentioning that researchers have used LSTM and its variants overwhelmingly with a view to forecasting COVID-19 cases in an early outbreak scenario [2, 4, 22]. Nevertheless, our analysis enlightens that CNN and multivariate CNN algorithms have performed really well compared to LSTM algorithm. A probable reason for this might be explained from [25]. Our dataset lacks large amount of data, whereas sufficient amount of data is preferred for LSTM algorithm to capture the essential features in time series data. For instance, the performance of LSTM is really laudable in terms of dealing with stock price data while historical data is available [20]. Sima and Akbar [23] showed that LSTM algorithm can be applied successfully for analysing business analytics data. On the other hand, CNN fundamentally aims at focusing on local features with more attention [27]. In spite of having, CNN has been really successful in extracting local features, patterns, trends and deviations. Moreover, CNN has successfully unearthed the complex nonlinear dependencies regarding the epidemic transmission. The number of daily death cases in Brazil, India, Russia and the UK have been projected with a forecasting horizon of 40 days with our best validated model CNN.

## 4. Conclusions

In this paper, four deep learning architectures: LSTM, GRU, CNN and multivariate CNN have been implemented with a view to forecasting new COVID-19 cases and new deaths in Brazil, India, Russia and the UK until late December 2020. We have used MSE as our loss function and used MAPE and nRMSE as the performance indicators for the studied deep learning models. Among the four deep learning models, CNN has performed extremely well in terms of validation accuracy and forecasting consistency. Moreover, the concept of multivariate CNN architecture has been implemented for the first time in this study, which showed robust forecasting performance for the studied countries. CNN algorithm has been proposed for long term forecasting in the absence of seasonality and periodic patterns in time series datasets. Moreover, the failure of LSTM architecture has also been disclosed in analysing such datasets. Importantly, according to CNN model projection results, clear indications pertinent to quick arrival of second wave of COVID-19 in above-mentioned countries have been unearthed. Due to pandemic fatigue and public apathy towards different non-pharmaceutical intervention strategies, second wave of COVID-19 could bring dreadful disasters in aforesaid countries. Public health officials in those countries should act immediately and deploy stringent intervention strategies to battle strategically against the pandemic.

## Data Availability

All datasets are available online

